# Evaluation of a Study Advisory Group on the Facilitators and Barriers to Patient Engagement

**DOI:** 10.64898/2026.11.16.26368203

**Authors:** Kirsten T. Schmidt, Sadia Ahmed, Fakhriyya Aghabayli, Kaniz Jannat, Brenda M.Y. Leung

## Abstract

The facilitators and barriers to patient engagement in an advisory group (AG) were evaluated. The AG was formed to provide the “patient” voice in the development of a pragmatic trial to study multinutrient treatment in children with ADHD. The AG consisted of patient partners (those with lived experience and caregivers) and healthcare professionals (those working with children with ADHD). Eleven AG members completed the Patient Engagement in Research Scale, and five members participated in semi-structured interviews. Qualitative data were analyzed thematically, and five themes were identified. Together with the quantitative findings, key facilitators of engagement included flexible engagement, a sense of trust among AG members and the research team, and meaningful opportunities to contribute. Barriers included irregular communication from the research team, ambiguity regarding members’ roles, and underutilization of members’ competencies. Findings highlight the importance of interpersonal relationships and team dynamics and informed improvements such as more regular communication, improved onboarding processes to strengthen role clarity, and better leveraging of members’ expertise to support full engagement. This evaluation provides insights into the use of research evaluation to strengthen patient engagement in applied research settings.

## 1.1 Background

Patient-centered priorities and outcomes are recognized as the cornerstones of patient-oriented research (POR), reflecting a shift from studying patients as research participants to collaborating with patients as research partners^1^. POR is a collaborative approach where patients and/or their caregivers, family members, friends, or community members with lived experience of specific health conditions help shape research decisions – from developing research questions to designing and conducting studies and translating outcomes about health issues that matter in everyday life^1^. This alignment increases the relevance of research findings, enhances the likelihood of results that can inform more meaningful and appropriate changes in healthcare policy and practice, and strengthens the real-world applicability of research results. The goal of patient engagement in research is to improve inclusivity and enhance trust and engagement throughout the research lifecycle.

Attention-Deficit/Hyperactivity Disorder (ADHD) is a neurodevelopmental disorder, and challenges to management and care continue to exist due to gaps in diagnosis and access to effective treatment and appropriate care^2^. Interest in research on nutritional interventions (e.g., micronutrients, polyunsaturated fatty acids, diet modifications) has grown, but studies often do not account for the lived realities, priorities, and constraints on children and caregivers managing ADHD^3^. Also, inclusive strategies for participant recruitment and retention remain ongoing challenges in clinical trials, particularly in populations experiencing complex health challenges and dynamic sociocultural and economic environments^4^ such as ADHD.

Recognizing the importance of conducting more patient-centered research and translation of findings to improve real-world outcomes, the Patient Oriented Innovative Nutrition Trial (POINT) study team sought to explore how patient and caregiver involvement could inform the design of an innovative clinical trial. The POINT study team established an advisory group (AG) of fourteen members with lived experience with ADHD or professionals with clinical, educational, or research expertise in ADHD to inform the design of a pragmatic trial. Using a patient-oriented approach to designing an innovative clinical trial, the POINT team consulted the AG to identify gaps informed by patients, thus integrating lived experience and expertise into their research design, processes, and dissemination. To better understand the facilitators and barriers to patient engagement, we conducted an evaluation of AG members in the POINT study on their perspectives and experiences of engaging in the project.

### 1.2 Formation of the Advisory Group

The POINT AG was formed in May 2024. Calls for members were made through the Alberta Strategy for Patient-Oriented Research (AbSPORU) Albertans4HealthResearch.ca network^5^, University of Lethbridge social media, and through the research teams’ own networks. The AG members identified mostly as female (12 out of 14) and represented diverse ethnic and racial backgrounds of West Asian, South Asian, East Asian, Black or African descent, and White. Members’ roles included Patient Partner, Family Partner, Health Professional, and Community support.

The AG was supported by the Research Coordinator (RC) and Research Assistant (RA), both of whom began as Patient Partners but evolved into these more comprehensive roles. Oversight was provided by the Principal Investigator (PI). The meeting schedule was decided by consensus, and meetings were held every third Friday of every third month via Zoom, an online video conference platform^6^. Members opted to have POINT RC chair the meetings. A co-chair position was offered, but no members volunteered. Agendas were developed by POINT RC and PI and distributed via email one week prior to every meeting. Notes were taken by the POINT RC during meetings and sent out via email for AG members to review after meetings, as well as uploaded to a shared University of Lethbridge OneDrive folder for all members to access.

A training session on patient-oriented research was delivered by the AbSPORU Patient Engagement Team, and AG members were encouraged to complete the online Patient-Oriented Research Curriculum in Child Health (PORCCH) modules^7^. Members were also invited to join smaller working groups that were established throughout the two-year term: Equity, Diversity, and Inclusion (EDI); Communications; and Training and Capacity Building. The EDI group focused on fostering an inclusive and equitable environment and ensuring accessible outputs; the Communications group supported internal and external communication, outreach efforts, and newsletters; and the Training and Capacity Building group identified and recommended training modules, helped coordinate meetings and two AG retreats. Other ad hoc working groups were formed on an as-needed basis. Compensation, according to AbSPORU patient partner compensation guidelines^8^, was offered to AG members for project related activities, such as completing training modules online, meeting attendance, being on working groups, reading and providing feedback on any project materials, and for participation in the patient engagement evaluation.

POINT AG members had the opportunity to be actively engaged in several collaborative activities throughout the project:

- ‘Terms of Reference’ and ‘Compensation Guidelines’ were co-written by the research team members (RA/RC/PI), reviewed by the POINT EDI working group, distributed via email to the AG members, and uploaded to the shared OneDrive for all members to access
- A POINT logo was created by the RA, then reviewed and changes suggested and approved by AG members
- Two one-day online retreats were hosted, during which patient partners collaboratively co-created several outputs including the *Collective Hope* infographic, the *ADHD and Nutrition Community of Practice* concept, the *ADHD Patient Journey Map* infographic, and a *Research Brief*
- AG members and the research team attended four conferences to network and present findings from the POINT project
- A rapid review of the literature was completed, and a manuscript was co-authored by the research team and a Rapid Review Working Group
- A mixed-methods patient engagement evaluation (as presented here) was conducted to inquire about AG member experience and their perspectives about what worked well and what aspects of the engagement processes could be changed/improved.

## 2.0 Methods

### 2.1 Evaluation Design

This evaluation collected quantitative and qualitative data using a standardized questionnaire and semi-structured interviews. The collection of both quantitative and qualitative data allowed both a structured assessment and in-depth exploration of engagement experiences, supporting a more comprehensive evaluation of the patient engagement processes. An AbSPORU Patient Engagement Team member (SA), who was not an AG member nor a member of the POINT research team, conducted the evaluation as an external evaluator. The study was approved by the University of Calgary Conjoint Health Research Ethics Board (REB20-1822). A presentation about the evaluation purpose, design, and methods was shared with the AG in February 2025 for feedback and questions. The online survey and one-on-one interviews were conducted mid-point through the POINT study, ten months after establishing the AG, via Zoom from March – April 2025.

### 2.2 Quantitative Survey and Qualitative Interview Guide

Through consultation with the research team members, the Patient Engagement in Research Scale (PEIRS-22), a validated evaluation tool^9,10^, was selected, and two open-ended questions were added. Other evaluation tools exist, including the Patient and Public Evaluation Engagement Tool (PPEET)^11^ and the Learning Together Evaluation Framework for Patient and Public Engagement^12^; however, the PEIRS tool was specifically co-developed and validated to evaluate meaningful experiences of patient partners on research teams^13^. The PEIRS-22 survey consisted of twenty-two items structured to the following domains: *procedural requirements, convenience, contributions, team environment and interaction, support, feel valued, and benefits*. Each item used a 5-point Likert scale that ranged from ‘strongly agree’ to ‘strongly disagree’. This informed an anonymous online survey hosted on the University of Calgary Qualtrics^14^ platform.

The interview guide (Appendix A) was co-developed by members of the POINT research team and the AbSPORU Patient Engagement Team. The open-ended questions in the interview guide were designed to allow participants to share deeper, more nuanced reflections on the specific activities they engaged in and their experiences working together as an AG. The interview guide also included specific questions on the support AG members received, feedback about communication strategies, and suggestions for ways the AG could improve.

### 2.3 Data Collection

All fourteen AG members were emailed the survey link on March 12, 2025. The survey remained open until April 2, 2025. The survey was conducted using Qualtrics^14^. The survey was anonymous, and no personal identifiers were collected. Members were also invited to participate in online Zoom interviews using the co-developed question guide. Interviews were conducted between April 9 and 29, 2025, and ranged between twenty-four to thirty-six minutes. Interviews were audio-recorded and transcribed verbatim.

### 2.4 Data Analysis

Survey responses were organized into a summary table and aggregated to report total counts and percentages. Likert-scale responses were grouped into agree (strongly agree/agree), neutral, and disagree (strongly disagree/disagree). Domain-level agreement percentages were calculated by averaging the proportion of respondents selecting “Agree/Strongly agree” across items within each of the seven domains.

Interview transcripts were uploaded to NVivo software^15^ and analyzed by SA using Braun & Clarke’s six-step method of reflective thematic analysis^16^. An inductive approach to coding was used, and codes were categorized into five main themes. Supporting quotes were identified to support these themes.

### 3.0 Results

Of the fourteen AG members, eleven (79%) completed the online survey and five (36%) participated in individual online interviews.

### 3.1 Quantitative data: PEIRS-22 Survey

PEIRS-22 survey results are reported in Table 1A. Overall, responses indicated positive perceptions of engagement across the seven domains.

**Table 1A.** Participant ratings of engagement quality across PEIRS-22 domains.

| Domains | Items | Agree<br>n (%) | Neutral<br>n (%) | Disagree<br>n (%) |
|---|---|---|---|---|
| **Procedural Requirements** |  |  |  |  |
|  | The research team members were properly introduced to each other | 7 (64%) | 3 (27%) | 0 (0%) |
|  | In general, I had sufficient opportunities to contribute to the project | 8 (73%) | 3 (27%) | 0 (0%) |
|  | I was able to perform my tasks for the project | 9 (82%) | 1 (9%) | 1 (9%) |
|  | I participated in making decisions about the project | 6 (55%) | 3 (27%) | 2 (18%) |
|  | I received sufficient updates about the project | 9 (82%) | 1 (9%) | 1 (9%) |
|  | Communication within the research team was clear throughout the project | 8 (73%) | 1 (9%) | 2 (18%) |
|  | The project was worth the time I spent on it | 8 (73%) | 3 (27%) | 0 (0%) |
| **Domain Summary** (mean % “Agree” across 7 items) = 72% |  |  |  |  |
| **Convenience** |  |  |  |  |
|  | I had the opportunity to provide input into selecting my tasks for the project | 9 (82%) | 0 (0%) | 2 (18%) |
|  | Throughout the project, I had sufficient time to complete my tasks for the project | 8 (73%) | 2 (18%) | 1 (9%) |
|  | I had opportunities to express my views | 11 (100%) | 0 (0%) | 0 (0%) |
| **Domain Summary** (mean % “Agree” across 3 items) = 85% |  |  |  |  |
| **Contributions** |  |  |  |  |
|  | I contributed by providing my perspective | 10 (91%) | 0 (0%) | 1 (9%) |
|  | My contributions were a good use of my time | 9 (82%) | 2 (18%) | 0 (0%) |
|  | My workload in the project was manageable | 10 (91%) | 1 (9%) | 0 (0%) |
| **Domain Summary** (mean % “Agree” across 3 items) = 88% |  |  |  |  |
| **Team Environment and Interaction** |  |  |  |  |
|  | I was an equal partner in the research project team | 4 (36%) | 5 (45%) | 2 (18%) |
|  | There was trust among the research project team members | 7 (64%) | 4 (36%) | 0 (0%) |
| **Domain Summary** (mean % “Agree” across 2 items) = 50% |  |  |  |  |
| **Support** |  |  |  |  |
|  | I received sufficient support to contribute to the project (for example, orientation, readings, training workshops, webinars) | 10 (91%) | 1 (9%) | 0 (0%) |
|  | Any concerns I had were addressed | 6 (55%) | 5 (45%) | 0 (0%) |
| **Domain Summary** (mean % “Agree” across 2 items) = 73% |  |  |  |  |
| **Feel Valued** |  |  |  |  |
|  | The research project team appreciated my contributions | 9 (82%) | 2 (18%) | 0 (0%) |
|  | I was offered sufficient recognition for my contributions (for example, payment, authorship, or gifts) | 9 (82%) | 1 (9%) | 1 (9%) |
| **Domain Summary** (mean % “Agree” across 2 items) = 82% |  |  |  |  |
| **Benefits** |  |  |  |  |
|  | I enjoyed being a part of the project | 9 (82%) | 2 (18%) | 0 (0%) |
|  | I made an impact on the decisions in the project | 6 (55%) | 2 (18%) | 3 (27%) |
|  | My involvement had positive impacts on my life | 6 (55%) | 3 (27%) | 2 (18%) |
| **Domain Summary** (mean % “Agree” across 3 items) = 64% |  |  |  |  |

Note: This table presents participant responses (n=11) to the PEIRS-22, summarized by domain. Survey Likert-scale responses were grouped into agree (strongly agree/agree), neutral, and disagree (strongly disagree/disagree). Responses are reported as frequencies and percentages for agreement, neutrality, and disagreement across individual items, along with mean percentage agreement for each domain. Domain-level agreement percentages were calculated by averaging the proportion of respondents selecting “Agree/Strongly Agree” across items within each domain.

#### Procedural Requirements

Between 55% (6/11) and 82% (9/11) of participants agreed that engagement processes were clear and well-structured. For example, 82% (9/11) agreed they were able to perform their project tasks, and 82% (9/11) felt they received sufficient updates. However, responses were more mixed regarding participation in decision-making, as only 55% (6/11) were in agreement.

#### Convenience

Convenience-related items were rated highly, with 73% (8/11) to 100% (11/11) agreement. Notably, all participants (100%) agreed they had opportunities to express their views.

#### Contributions

High levels of agreement were observed regarding participants’ contributions, with ratings of agreement between 82% (9/11) and 91% (10/11). Most respondents felt their contributions were a good use of time 82% (9/11) and that their workload was manageable 91% (10/11).

#### Team Environment and Interaction

Perceptions of team dynamics were generally positive, with 64% (7/11) agreeing that trust existed among team members. However, only 36% (4/11) agreed they were equal partners, with 45% (5/11) selecting neutral.

#### Support

Support-related items were rated positively, with 91% (10/11) agreeing they received sufficient support and 55% (6/11) agreeing that concerns were addressed. The remaining 45% (5/11) voted neutral.

#### Feeling Valued

Most participants felt valued within the project, with 82% (9/11) agreeing that their contributions were appreciated and recognized.

#### Benefits

While 82% (9/11) reported enjoying their collaboration, perceptions of broader impact were more variable. Only 55% (6/11) agreed they made an impact on project decisions, and 55% (6/11) agreed that involvement had positive impacts on their life, with some disagreement noted 18-27%.

### 3.2 Open-ended survey questions

The survey’s first open-ended question, “What are the strengths of the POINT Advisory Group?” received positive feedback from eight survey respondents (Table 1B). Overall, members felt “*the meetings were dynamic,*” with “*representation of diverse experiences*” and the “*many perspectives*” involved in a single topic. The research team was attributed with doing a good job of making members feel “*welcomed and included*.” Participants shared that the working groups supported members to “*choose to be more or less involved depending on [their] capacity and free time*,”. Working groups were referred to as “*opportunities to participate*” and “*ways to engage parent-partners*.” Participants also highlighted the value of these working groups in offering flexibility in engagement, noting appreciation for being able to “*choose to be more or less involved depending on [their] capacity and free time.*”

**Table 1B.** PEIRS-22 survey open-ended responses.

| Question | Participant Responses |
|---|---|
| What are the strengths of the POINT Advisory Group? (n=8) | ● I like how I was introduced to the whole team, the meetings were dynamic and kept my interest, the ways each member connected with me. [The POINT Research Assistant] really made me feel welcomed and included. I liked the interactive portions she led<br>● It's well organized, easy to follow instructions and easy and well communicated.<br>● Draw on experience and expertise outside academia<br>● Education around project, giving sufficient information on background (including literature and rapid review info)<br>● Representation of diverse experiences<br>● I like the diversity of people involved into the Advisory Group at POINT. It gives many perspectives at the same topics and is a great opportunity to learn from it too.<br>● lots of ways to engage parent-partners<br>● Lots of opportunities to participate<br>● I like how I can choose to be more or less involved depending on my capacity and free time |
| What can we improve on in the POINT Advisory Group? (n=6) | ● Perhaps consider using streamlines project management tools, there are resources distributed through shared folders, emails etc, a central project management tool may help to enhance the experience.<br>● Continuous recruiting<br>● Communicating specific tasks of the working groups to help guide whether joining would be something I could do or have the capacity for, recognizing barriers to participation/allowing for more breaks during longer sessions, usually receive gift cards for honorarium but this was discouraged and not accepted as alternative option<br>● More frequent and clear and timely communication<br>● More explanations about the MADDY 2.0 project<br>● Keep the goals of the study in mind. Sometimes, focus shifts to general ADHD advocacy, which is interesting, but not related to the study. |

Note: This table presents participant responses to the open-ended questions that were added to the PEIRS-22 regarding strengths and areas for improvement of the POINT Advisory Group. Responses are presented verbatim and are grouped by survey questions.

The survey’s second open-ended question, “What can we improve on in the POINT Advisory Group?” received responses from six survey participants (Table 1B). Although the POINT project utilizes the University of Lethbridge Microsoft system, difficulties have been encountered as members find it difficult to access/overcome the firmware. Members commented on the need for a more accessible “*central project management tool may help to enhance the experience*.”

On a similar topic, a member shared that “*communicating specific tasks of the working groups to help guide whether joining would be something I could do or have the capacity for.*” A suggestion during a post-evaluation discussion suggested that a list of working groups and their outputs, goals, and timelines could help.

Finally, some members mentioned that they would have liked to receive more plain-language explanations and analyses of the previous Micronutrients for ADHD in Youth (MADDY) project/results ^17^. Without clear guiding background information, the discussions and “*focus often shift[ed] to general ADHD advocacy.*” If the research team could guide conversations regarding needs for the clinical trial, they would be better able to provide relevant and meaningful input.

### 3.3 Qualitative data: Semi-structured interviews

Five main themes resulted from the thematic analysis of the semi-structured interview data: (1) Project-focused communication, (2) working with group dynamics while respecting diversity of members, (3) fostering active and meaningful involvement, (4) what’s working well in POINT, and (5) member hopes and goals for POINT. These themes, along with supportive quotes, are shared in Table 2.

**Table 2.**
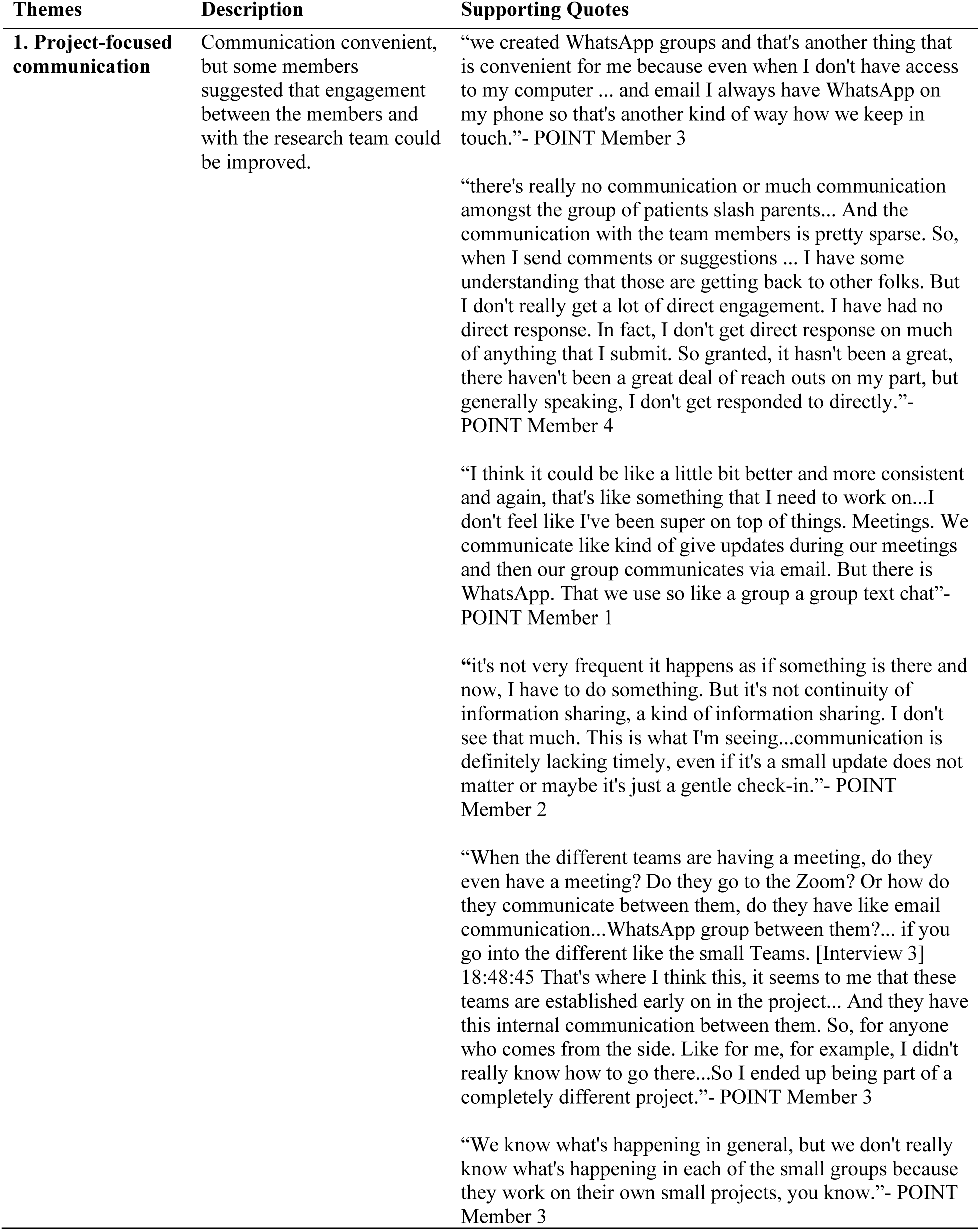
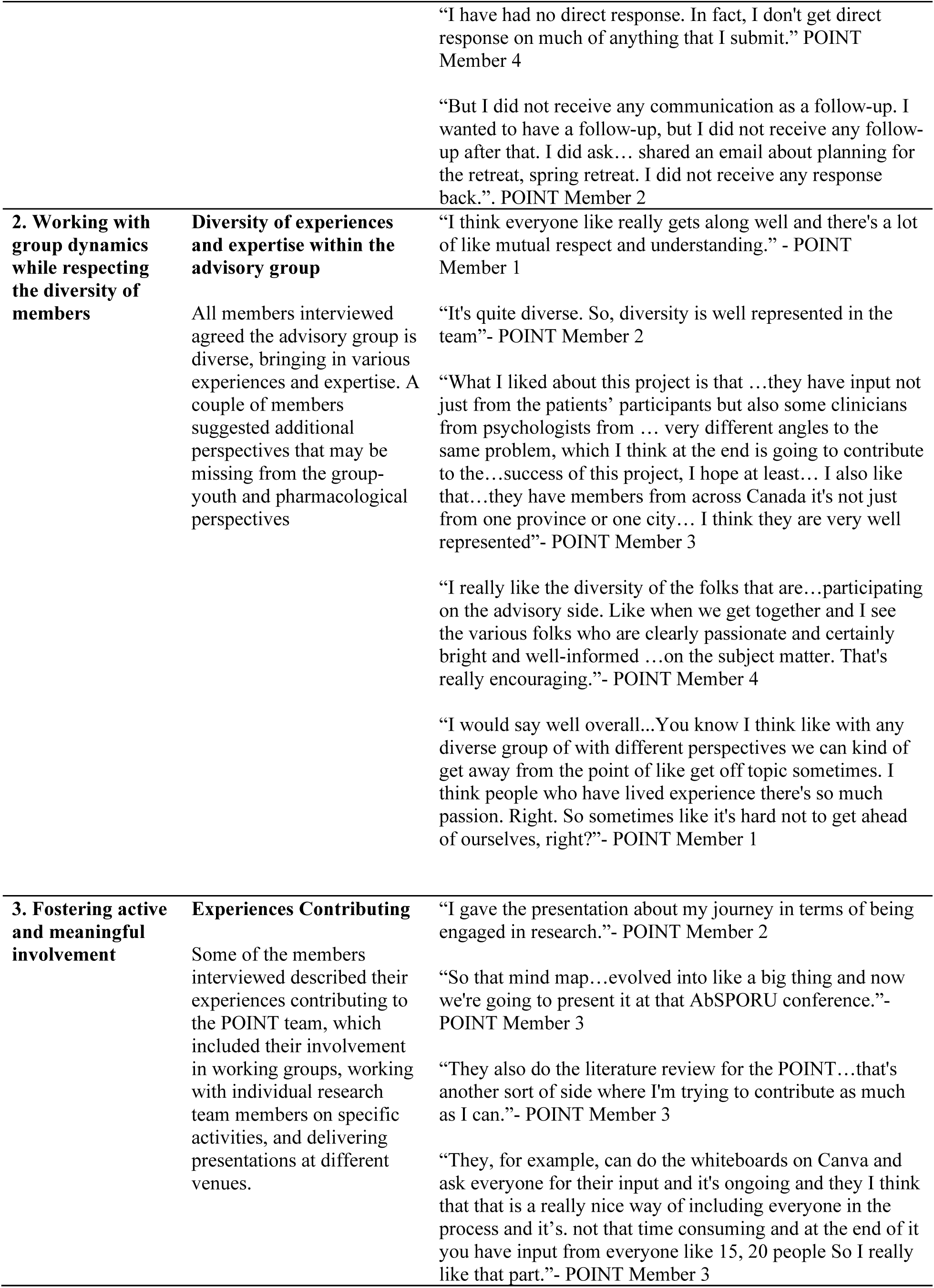
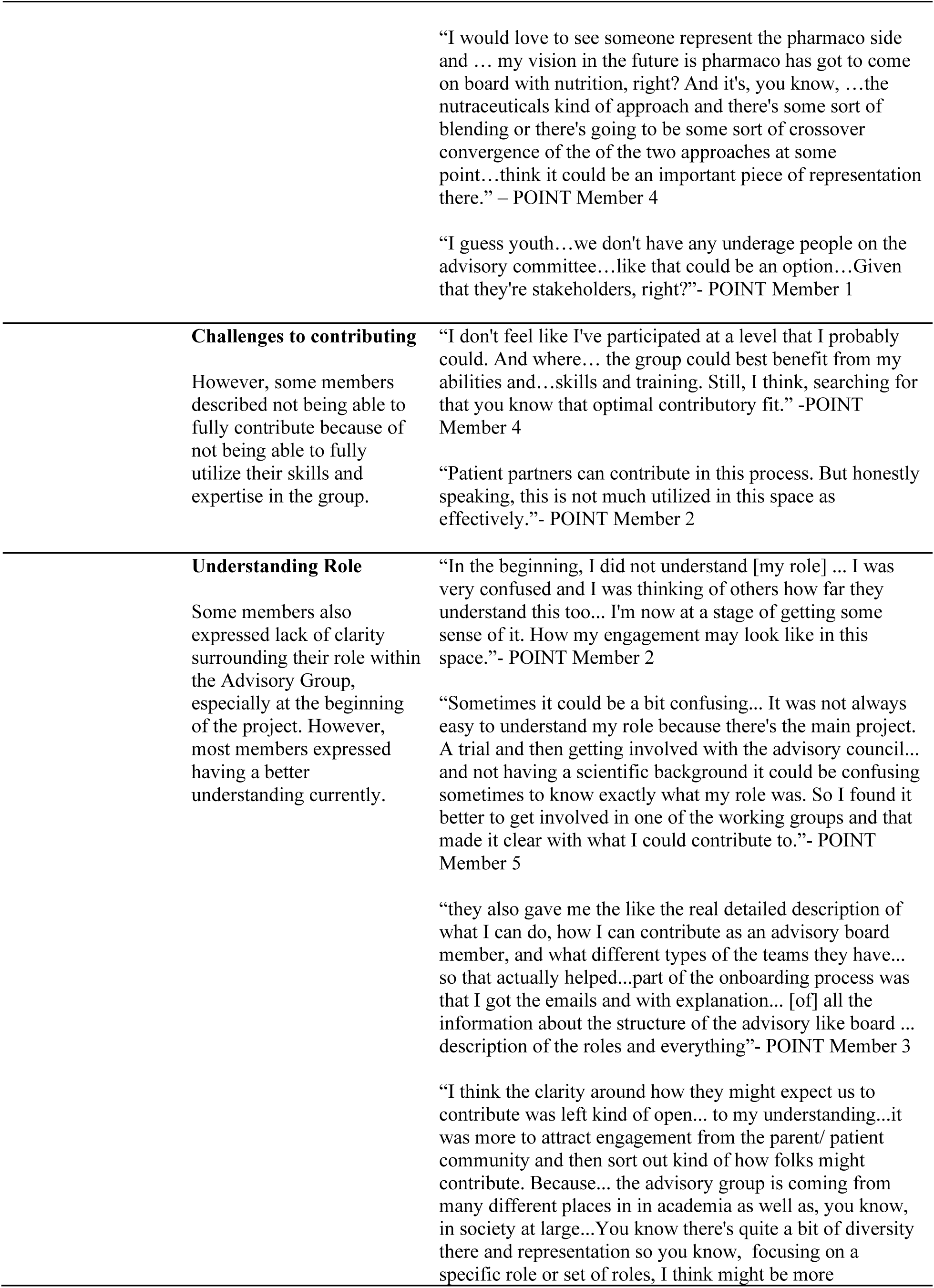
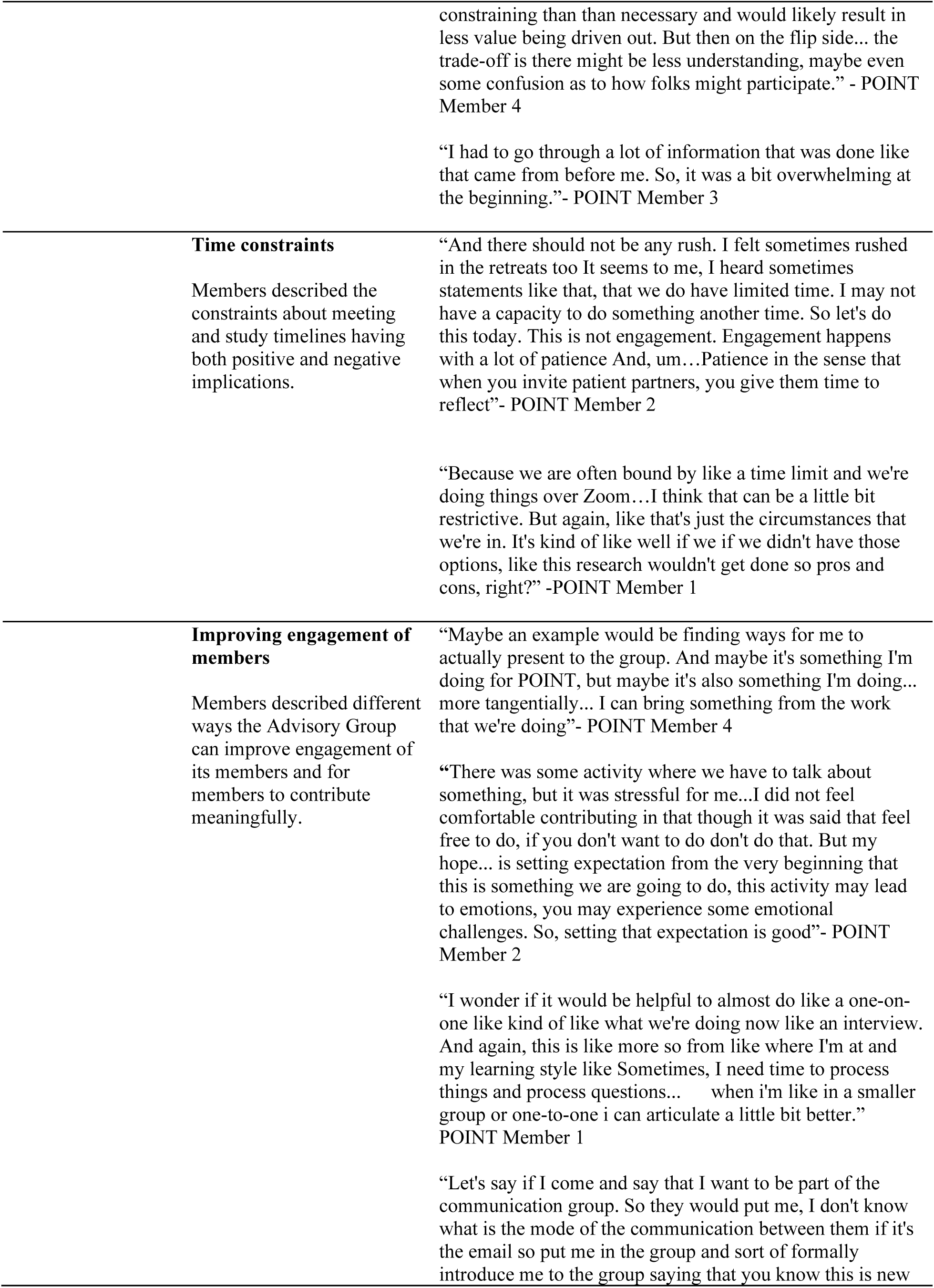
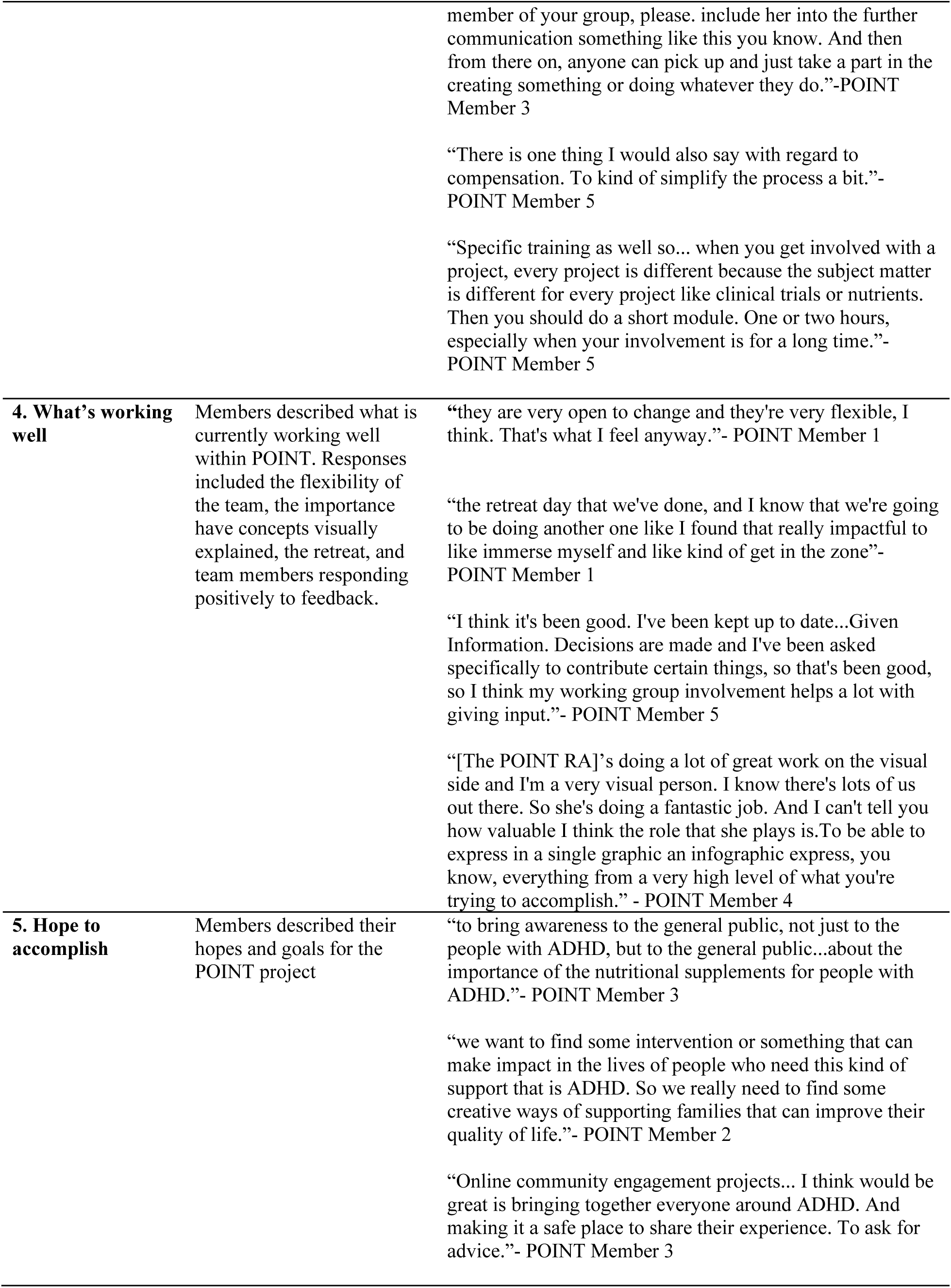
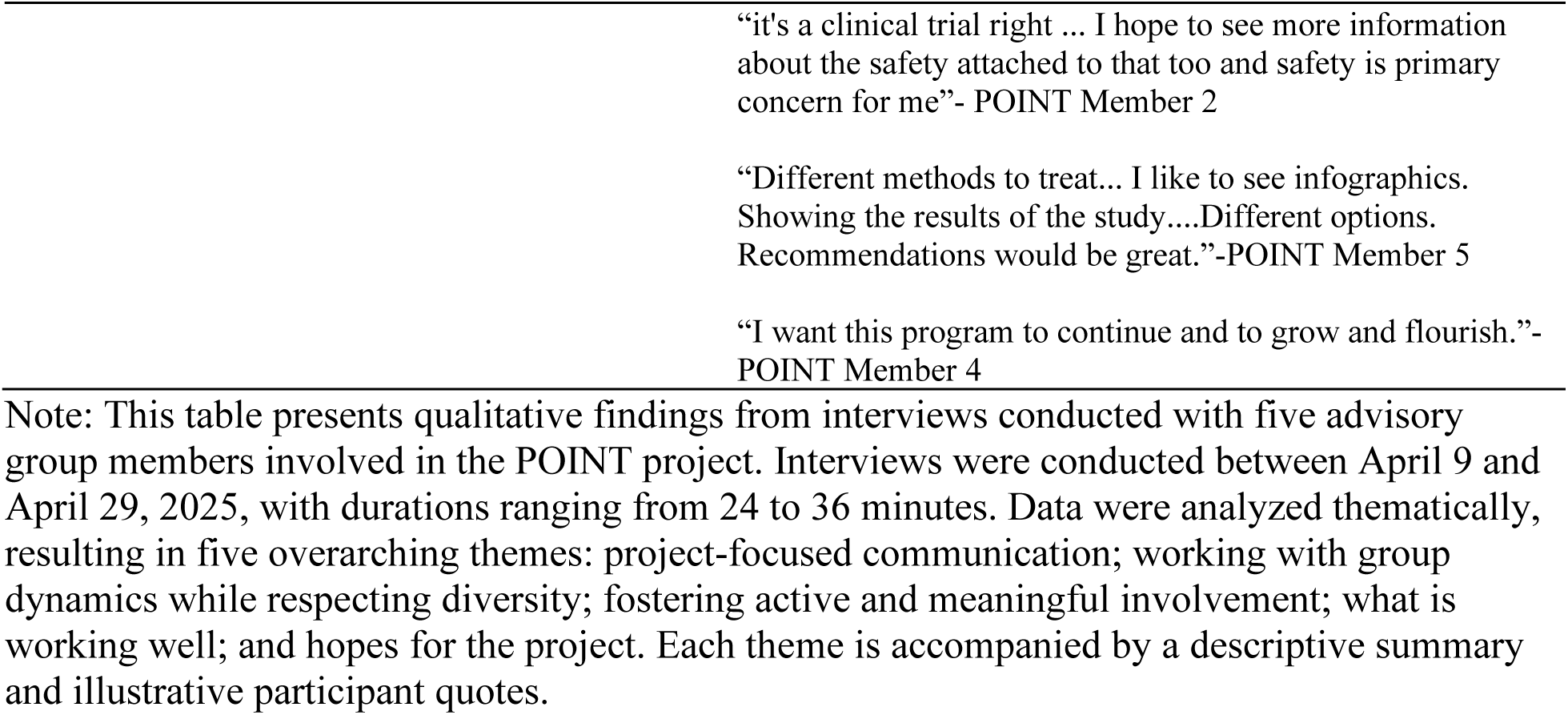
Thematic findings from semi-structured interviews of advisory group members.

| Themes | Description | Supporting Quotes |
|---|---|---|
| **1. Project-focused communication** | Communication convenient, but some members suggested that engagement between the members and with the research team could be improved. | “we created WhatsApp groups and that's another thing that is convenient for me because even when I don't have access to my computer ... and email I always have WhatsApp on my phone so that's another kind of way how we keep in touch.”- POINT Member 3<br>“there's really no communication or much communication amongst the group of patients slash parents... And the communication with the team members is pretty sparse. So, when I send comments or suggestions ... I have some understanding that those are getting back to other folks. But I don't really get a lot of direct engagement. I have had no direct response. In fact, I don't get direct response on much of anything that I submit. So granted, it hasn't been a great, there haven't been a great deal of reach outs on my part, but generally speaking, I don't get responded to directly.”- POINT Member 4<br>“I think it could be like a little bit better and more consistent and again, that's like something that I need to work on...I don't feel like I've been super on top of things. Meetings. We communicate like kind of give updates during our meetings and then our group communicates via email. But there is WhatsApp. That we use so like a group a group text chat”- POINT Member 1<br>**“**it's not very frequent it happens as if something is there and now, I have to do something. But it's not continuity of information sharing, a kind of information sharing. I don't see that much. This is what I'm seeing...communication is definitely lacking timely, even if it's a small update does not matter or maybe it's just a gentle check-in.”- POINT Member 2<br>“When the different teams are having a meeting, do they even have a meeting? Do they go to the Zoom? Or how do they communicate between them, do they have like email communication...WhatsApp group between them?... if you go into the different like the small Teams. [Interview 3] 18:48:45 That's where I think this, it seems to me that these teams are established early on in the project... And they have this internal communication between them. So, for anyone who comes from the side. Like for me, for example, I didn't really know how to go there...So I ended up being part of a completely different project.”- POINT Member 3<br>“We know what's happening in general, but we don't really know what's happening in each of the small groups because they work on their own small projects, you know.”- POINT Member 3<br>“I have had no direct response. In fact, I don't get direct response on much of anything that I submit.” POINT Member 4<br>“But I did not receive any communication as a follow-up. I wanted to have a follow-up, but I did not receive any follow-up after that. I did ask… shared an email about planning for the retreat, spring retreat. I did not receive any response back.”. POINT Member 2 |
| **2. Working with group dynamics while respecting the diversity of members** | **Diversity of experiences and expertise within the advisory group**<br>All members interviewed agreed the advisory group is diverse, bringing in various experiences and expertise. A couple of members suggested additional perspectives that may be missing from the group-youth and pharmacological perspectives | “I think everyone like really gets along well and there's a lot of like mutual respect and understanding.” - POINT Member 1<br>“It's quite diverse. So, diversity is well represented in the team”- POINT Member 2<br>“What I liked about this project is that …they have input not just from the patients’ participants but also some clinicians from psychologists from … very different angles to the same problem, which I think at the end is going to contribute to the…success of this project, I hope at least… I also like that…they have members from across Canada it's not just from one province or one city… I think they are very well represented”- POINT Member 3<br>“I really like the diversity of the folks that are…participating on the advisory side. Like when we get together and I see the various folks who are clearly passionate and certainly bright and well-informed …on the subject matter. That's really encouraging.”- POINT Member 4<br>“I would say well overall...You know I think like with any diverse group of with different perspectives we can kind of get away from the point of like get off topic sometimes. I think people who have lived experience there's so much passion. Right. So sometimes like it's hard not to get ahead of ourselves, right?”- POINT Member 1 |
| **3. Fostering active and meaningful involvement** | **Experiences Contributing**<br>Some of the members interviewed described their experiences contributing to the POINT team, which included their involvement in working groups, working with individual research team members on specific activities, and delivering presentations at different venues. | “I gave the presentation about my journey in terms of being engaged in research.”- POINT Member 2<br>“So that mind map…evolved into like a big thing and now we're going to present it at that AbSPORU conference.”- POINT Member 3<br>“They also do the literature review for the POINT…that's another sort of side where I'm trying to contribute as much as I can.”- POINT Member 3<br>“They, for example, can do the whiteboards on Canva and ask everyone for their input and it's ongoing and they I think that that is a really nice way of including everyone in the process and it’s. not that time consuming and at the end of it you have input from everyone like 15, 20 people So I really like that part.”- POINT Member 3<br>“I would love to see someone represent the pharmaco side and … my vision in the future is pharmaco has got to come on board with nutrition, right? And it's, you know, …the nutraceuticals kind of approach and there's some sort of blending or there's going to be some sort of crossover convergence of the of the two approaches at some point…think it could be an important piece of representation there.” – POINT Member 4<br>“I guess youth…we don't have any underage people on the advisory committee…like that could be an option…Given that they're stakeholders, right?”- POINT Member 1 |
|  | **Challenges to contributing**<br>However, some members described not being able to fully contribute because of not being able to fully utilize their skills and expertise in the group. | “I don't feel like I've participated at a level that I probably could. And where… the group could best benefit from my abilities and…skills and training. Still, I think, searching for that you know that optimal contributory fit.” -POINT Member 4<br>“Patient partners can contribute in this process. But honestly speaking, this is not much utilized in this space as effectively.”- POINT Member 2 |
|  | **Understanding Role**<br>Some members also expressed lack of clarity surrounding their role within the Advisory Group, especially at the beginning of the project. However, most members expressed having a better understanding currently. | “In the beginning, I did not understand [my role] ... I was very confused and I was thinking of others how far they understand this too... I'm now at a stage of getting some sense of it. How my engagement may look like in this space.”- POINT Member 2<br>“Sometimes it could be a bit confusing... It was not always easy to understand my role because there's the main project. A trial and then getting involved with the advisory council... and not having a scientific background it could be confusing sometimes to know exactly what my role was. So I found it better to get involved in one of the working groups and that made it clear with what I could contribute to.”- POINT Member 5<br>“they also gave me the like the real detailed description of what I can do, how I can contribute as an advisory board member, and what different types of the teams they have... so that actually helped...part of the onboarding process was that I got the emails and with explanation... [of] all the information about the structure of the advisory like board ... description of the roles and everything”- POINT Member 3<br>“I think the clarity around how they might expect us to contribute was left kind of open... to my understanding...it was more to attract engagement from the parent/ patient community and then sort out kind of how folks might contribute. Because... the advisory group is coming from many different places in in academia as well as, you know, in society at large...You know there's quite a bit of diversity there and representation so you know, focusing on a specific role or set of roles, I think might be more constraining than than necessary and would likely result in less value being driven out. But then on the flip side... the trade-off is there might be less understanding, maybe even some confusion as to how folks might participate.” - POINT Member 4<br>“I had to go through a lot of information that was done like that came from before me. So, it was a bit overwhelming at the beginning.”- POINT Member 3 |
|  | **Time constraints**<br>Members described the constraints about meeting and study timelines having both positive and negative implications. | “And there should not be any rush. I felt sometimes rushed in the retreats too It seems to me, I heard sometimes statements like that, that we do have limited time. I may not have a capacity to do something another time. So let's do this today. This is not engagement. Engagement happens with a lot of patience And, um…Patience in the sense that when you invite patient partners, you give them time to reflect”- POINT Member 2<br>“Because we are often bound by like a time limit and we're doing things over Zoom…I think that can be a little bit restrictive. But again, like that's just the circumstances that we're in. It's kind of like well if we if we didn't have those options, like this research wouldn't get done so pros and cons, right?” -POINT Member 1 |
|  | **Improving engagement of members**<br>Members described different ways the Advisory Group can improve engagement of its members and for members to contribute meaningfully. | “Maybe an example would be finding ways for me to actually present to the group. And maybe it's something I'm doing for POINT, but maybe it's also something I'm doing... more tangentially... I can bring something from the work that we're doing”- POINT Member 4<br>**“**There was some activity where we have to talk about something, but it was stressful for me...I did not feel comfortable contributing in that though it was said that feel free to do, if you don't want to do don't do that. But my hope... is setting expectation from the very beginning that this is something we are going to do, this activity may lead to emotions, you may experience some emotional challenges. So, setting that expectation is good”- POINT Member 2<br>“I wonder if it would be helpful to almost do like a one-on-one like kind of like what we're doing now like an interview. And again, this is like more so from like where I'm at and my learning style like Sometimes, I need time to process things and process questions... when i'm like in a smaller group or one-to-one i can articulate a little bit better.” POINT Member 1<br>“Let's say if I come and say that I want to be part of the communication group. So they would put me, I don't know what is the mode of the communication between them if it's the email so put me in the group and sort of formally introduce me to the group saying that you know this is new member of your group, please. include her into the further communication something like this you know. And then from there on, anyone can pick up and just take a part in the creating something or doing whatever they do.”-POINT Member 3<br>“There is one thing I would also say with regard to compensation. To kind of simplify the process a bit.”- POINT Member 5<br>“Specific training as well so... when you get involved with a project, every project is different because the subject matter is different for every project like clinical trials or nutrients. Then you should do a short module. One or two hours, especially when your involvement is for a long time.”- POINT Member 5 |
| **4. What’s working well** | Members described what is currently working well within POINT. Responses included the flexibility of the team, the importance have concepts visually explained, the retreat, and team members responding positively to feedback. | **“**they are very open to change and they're very flexible, I think. That's what I feel anyway.”- POINT Member 1<br>“the retreat day that we've done, and I know that we're going to be doing another one like I found that really impactful to like immerse myself and like kind of get in the zone”- POINT Member 1<br>“I think it's been good. I've been kept up to date...Given Information. Decisions are made and I've been asked specifically to contribute certain things, so that's been good, so I think my working group involvement helps a lot with giving input.”- POINT Member 5<br>“[The POINT RA]’s doing a lot of great work on the visual side and I'm a very visual person. I know there's lots of us out there. So she's doing a fantastic job. And I can't tell you how valuable I think the role that she plays is.To be able to express in a single graphic an infographic express, you know, everything from a very high level of what you're trying to accomplish.” - POINT Member 4 |
| **5. Hope to accomplish** | Members described their hopes and goals for the POINT project | “to bring awareness to the general public, not just to the people with ADHD, but to the general public...about the importance of the nutritional supplements for people with ADHD.”- POINT Member 3<br>“we want to find some intervention or something that can make impact in the lives of people who need this kind of support that is ADHD. So we really need to find some creative ways of supporting families that can improve their quality of life.”- POINT Member 2<br>“Online community engagement projects... I think would be great is bringing together everyone around ADHD. And making it a safe place to share their experience. To ask for advice.”- POINT Member 3<br>“it's a clinical trial right ... I hope to see more information about the safety attached to that too and safety is primary concern for me”- POINT Member 2<br>“Different methods to treat... I like to see infographics. Showing the results of the study....Different options. Recommendations would be great.”-POINT Member 5<br>“I want this program to continue and to grow and flourish.”- POINT Member 4 |

Note: This table presents qualitative findings from interviews conducted with five advisory group members involved in the POINT project. Interviews were conducted between April 9 and April 29, 2025, with durations ranging from 24 to 36 minutes. Data were analyzed thematically, resulting in five overarching themes: project-focused communication; working with group dynamics while respecting diversity; fostering active and meaningful involvement; what is working well; and hopes for the project. Each theme is accompanied by a descriptive summary and illustrative participant quotes.

**Table 3.** Recommendations for future engagement activities in response to study findings on facilitators and barriers.

| **Category** | **Barriers or Facilitators** | **Description (Evidence)** | **Recommendations** |
|---|---|---|---|
| Project-focused communication | Facilitators | Communication tools (WhatsApp, Zoom) were viewed as convenient and accessible. | Continue using multi-platform communication; maintain low-barrier tools for quick updates. |
|  | Barriers | Communication within groups seen as “sparse,” “indirect,” and lacking continuity; little insight into working-group activities. | Increase frequency of updates; implement a brief monthly summary; share working-group updates or shared progress tracker. |
| Working with group dynamics while respecting diversity of members | Facilitators | A diverse group with high level of trust, mutual respect, and appreciation; members felt welcomed and supported. | Maintain inclusive culture; continue encouraging varied levels of engagement to match capacity. |
|  | Barriers | Limited representation of youth and pharmacological perspectives; challenges maintaining focused discussion in larger group formats. | Intentional recruitment of youth and pharmacological expertise, and use structured facilitation strategies (e.g., smaller breakout groups) to maintain focus. |
| Fostering Active and Meaningful Involvement | Facilitators | Clear onboarding and role descriptions; multiple, flexible pathways for contribution; smaller working groups and inclusive tools that support meaningful engagement across diverse skills and comfort levels. | Continue offering varied engagement formats; keep providing background materials for those who want deeper involvement. |
|  | Barriers | Initial role ambiguity and information overload; time-limited, large-group, and virtual formats that sometimes constrained reflection and meaningful contribution. | Strengthen onboarding packages; develop simplified role descriptions. Increase co-decision opportunities; incorporate more retreats or hybrid options when possible. |
| What’s Working Well in POINT | Facilitators | Flexibility; clear instructions; diverse experiences represented; retreats especially valued. | Maintain flexible engagement models; continue retreats; preserve use of visuals in communication outputs. |
| Member Hopes and Goals for POINT | Barriers | Difficulty accessing the University Microsoft system; limited compensation options; need clearer trial background and plain-language summaries. | Adopt more accessible project management tools; expand compensation options (e.g., gift cards); provide ongoing plain-language trial updates, infographics, and safety information. |

Note: This table presents identified barriers and facilitators to advisory group engagement, organized by themes derived from qualitative interviews with five advisory group members in the POINT project. For each theme, barriers and facilitators are summarized alongside supporting evidence and corresponding recommendations to enhance engagement and participation.

#### 3.3.1 Theme 1: Project-focused communication

Communication between the research team and AG members was identified as the most challenging aspect of the process; several participants reported it as a modifiable barrier. For example, one member voiced minimal communication between the research team and AG members, noting: *“There’s really no communication or much communication amongst the group of patients/ parents… And the communication with the team members is pretty sparse”,* POINT Member 4. Another member stated: *“It’s not very frequent; it happens as if something is there and now, I have to do something”,* POINT member 2.

In addition to the frequency of communication, one member pointed out a lack of communication on the activities of smaller working groups and opportunities for other group members, *“We know what’s happening in general, but we don’t really know what’s happening in each of the small groups because they work on their own small projects, you know……I didn’t really know how to go there……”,* POINT member 3.

Participants also shared their frustration with not getting any response from the research team on time. POINT member 4 mentioned, *“I have had no direct response. In fact, I don’t get direct response on much of anything that I submit.”* POINT member 2 also echoed this observation, mentioning, *“But I did not receive any communication as a follow-up. I wanted to have a follow-up, but I did not receive any follow-up after that. I did ask… shared an email about planning for the retreat, spring retreat. I did not receive any response back.”*

#### 3.3.2 Theme 2: Working with group dynamics while respecting the diversity of members

A number of participants expressed appreciation for the group dynamics, particularly the diversity and richness of perspectives within the group. One member described the group as positive and supportive: *“I think everyone like really gets along well, and there’s a lot of like mutual respect and and understanding […]”,* POINT member 1. While some of the AG members highlighted the range of perspectives that each member brought to the team. Another person appreciated having a representative across Canada, “*I also like that…they have members from across Canada; it’s not just from one province or one city… I think they are very well represented*”, POINT member 3. Another member stated: “*When we get together, and I see the various folks who are clearly passionate and certainly bright and well-informed…on the subject matter. That’s really encouraging*”, POINT member 4. The only challenge identified in relation to the range of perspectives was that discussions occasionally “*get off topic sometimes*”, POINT member 1. However, this was attributed to the enthusiasm associated with lived experience: “*I think people who have lived experience, there’s so much passion. Right. So sometimes, it’s hard not to get ahead of ourselves, right?* POINT member 1.

#### 3.3.3 Theme 3: Fostering active and meaningful involvement

Participants expressed mixed views regarding their involvement as AG members. Some described their participation as meaningful and active, whereas others highlighted aspects of the experience that could be strengthened. Members described several ways they contributed to the POINT advisory group, which included their involvement in working groups, working with individual research team members on specific activities, and delivering presentations at different venues. For example, a member described presenting their lived experience in research engagement, explaining that they “*Gave the presentation about [their] journey in terms of being engaged in research*”, POINT Member 2.

Others described contributing to project output and knowledge dissemination, including developing materials that later evolved into conference presentations. As a member explained, a brainstorming activity “*Evolved into a big thing and now we’re going to present it at that AbSPORU conference*”, POINT Member 3. Members also described contributing to research activities such as literature reviews and collaborative brainstorming. One participant noted that interactive tools, such as shared virtual whiteboards, helped facilitate inclusive input from many members, describing it as “*A really nice way of including everyone in the process, it’s not that time-consuming, and at the end of it you have input from everyone, like 15, 20 people*”, POINT Member 3.

Despite these opportunities, some members felt they had not yet been able to fully contribute their skills or expertise. Member 4 reflected that they had not been able to use their full potential, stating: *“Participated at a level that I probably could… where the group could best benefit from my abilities and skills*,” describing an ongoing effort to find the “*optimal contributory fit*” within the advisory group. Similarly, POINT Member 2 echoed this concern, stating: *“Patient partners can contribute to this process. But honestly speaking, this is not much utilized in this space as effectively.”* Additionally, engagement as an AG member was described as unclear to some of the participants. Some felt initial uncertainty regarding their role, particularly during the early stages of engagement. POINT Member 2 explained that “*In the beginning, I did not understand [my role]… I was very confused. I’m now at a stage of getting some sense of it. How my engagement may look like in this space.*”

Another participant (POINT Member 5) highlighted their scientific capacity and experience, *“Sometimes it could be a bit confusing… It was not always easy to understand my role… and not having a scientific background, it could be confusing sometimes to know exactly what my role was”*.

A few members suggested that additional perspectives may strengthen the advisory group, particularly youth and pharmacological expertise. One member highlighted the potential value of including a pharmacological perspective, noting the importance of greater collaboration between medication and nutrition approaches: “*I would love to see someone represent the pharmaco side…my vision in the future is pharmaco has got to come on board with nutrition…there’s going to be some sort of crossover or convergence of the two approaches at some point*”, POINT member 4.

Another member suggested including youth representation, noting that the advisory committee currently does not include younger voices despite their role as key stakeholders, “*I guess youth…we don’t have any underage people on the advisory committee…like that could be an option…given that they’re stakeholders, right?*”, POINT Member 1.

#### 3.3.4 Theme 4: What’s working well in POINT

Regarding engagement and collaboration, the retreats were highlighted as integral to facilitating their contribution, “*The retreat day that we’ve done, and I know that we’re going to be doing another one, like I found that really impactful to like immerse myself and like kind of get in the zone*”, POINT member 1. Members also appreciated the POINT RA’s digital design skills as she supported the visualization of many POINT outputs using illustrative tools, “*And I can’t tell you how valuable I think the role that she plays is. To be able to express in a single graphic […] everything from a very high level of what you’re trying to accomplish*”, POINT member 4.

One member summarized their overall experience as positive, “*I think it’s been good. I’ve been kept up to date…Given Information. Decisions are made, and I’ve been asked specifically to contribute certain things, so that’s been good, so I think my working group involvement helps a lot with giving input*”, POINT member 5.

#### 3.3.5 Theme 5: Member hopes and goals for POINT

Many members shared their hopes and goals, both long-term and short-term, in the POINT study. Participants discussed opportunities for the project to increase public awareness and provide additional support for individuals and families affected by ADHD. One member described the project as an opportunity, “*To bring awareness to the general public […] about the importance of the nutritional supplements for people with ADHD*”, POINT member 3. Another emphasized the need for creative ways to support families, noting the importance of identifying interventions that could, “*Make [an] impact in the lives of people who need this kind of support*”, POINT Member 2. Members suggested that an online community could be a way to facilitate this support, describing it as a way of, “*Bringing together everyone around ADHD and making it a safe place to share their experience [and] to ask for advice*”, POINT member 3.

In addition, members highlighted the importance of providing clear and accessible information about the study and potential treatment approaches. For example, one participant emphasized that because the project involves a clinical trial, “*safety is [a] primary concern*” and expressed interest in seeing more information about the safety of interventions, POINT member 2. Another suggested using accessible formats, such as infographics, to communicate results and outline “*Different methods to treat…[and] different options [or] recommendations*”, POINT member 5.

### 4.0 Discussion

This evaluation provided valuable insights into the patient engagement experiences of members of the AG for the POINT project. Our findings illustrate both the benefits and complexities of integrating people with lived experience into research processes. Our evaluation revealed numerous facilitators that supported meaningful engagement, such as group flexibility, diversity of perspectives, welcoming culture, and a sense of comfort and trust within the team. However, some members described several challenges that impeded participation: infrequent communication, ambiguity in roles, responsibilities, or expectations, and underutilization of skills and expertise. Our study findings are consistent with previous findings that clarity of roles, accessible communication, and flexible approaches to engagement are essential for sustaining equitable collaboration^18^; our results reinforce these principles while adding context-specific insights from the POINT project. The positive relational factors that the AG members pointed out mirror findings in the engagement literature that emphasize the importance of psychological safety, mutual respect, and opportunities to contribute in ways that match personal capacity^18^. As identified by previous research, our study highlighted the effects of inclusivity of all patient-partners as a facilitator of this study^19^.

In addition, our evaluation reinforces prior studies that highlight that clarity of roles, accessible communication, and flexible approaches to engagement are essential for sustaining equitable collaboration^20^. Although regular communication with AG members was maintained throughout the study, members perceived it as insufficient, highlighting a gap between AG members’ expectations and the research team’s understanding.

Most importantly, the evaluation itself helped catalyze improvements within the POINT study itself. Member contributions strengthened the processes and the products of the research, reinforcing evidence that co-designing studies with patient partners enhances relevance, feasibility, and impact. In direct response to member feedback, the research team implemented several changes: a concise handout outlining roles, tasks, and timelines for the working groups and upcoming POINT outputs; individualized trouble-shooting sessions and guidance sessions for logistical support; and intentional “open space” in meetings to allow members to share their lived experiences and input on next steps. These modifications reflect the value of continuous quality improvement in patient engagement and support recent calls for adaptive and responsive engagement processes.

A key strength of this evaluation is the collection of both quantitative and qualitative data to give a deeper understanding of the patient partner experience by integrating survey findings and interview perspectives. The ongoing involvement of the research team and AG members contributed to the validity and authenticity of this evaluation, ensuring the interpretations reflected the group’s experiences. Additionally, the diversity within the AG – across geography, experience and perspectives – further enriched the insights generated. The one-year project extension represented an additional strength, as the team was able to complete POINT study outputs more thoughtfully and created additional opportunities for co-creation, including virtual retreats. Our team was open to feedback and incorporated what we could for the remaining project duration to improve relationships and engagement with the AG.

A limitation of this study was that the evaluation occurred at a single mid-project time point, which prevented us from examining changes over time or assessing the impact of the implemented improvements. A follow-up or exit survey could help address this gap. Additionally, the small sample size for both the survey and the interviews limited the breadth of insights and experiences in our findings.

### 4.1 Implications for Research Evaluation Practices

This study offers several implications for evaluation of patient engagement in research-focused patient advisory groups. Our findings highlight important challenges in the application and interpretation of engagement measures. This evaluation reinforces the importance of considering interpersonal relationships and team dynamics. While interpersonal relationships such as trust, respect, and inclusivity facilitated meaningful involvement, suboptimal team dynamics such as infrequent communication, ambiguity in roles, responsibilities, or expectations, and underutilization of skills and expertise limited participation. Effective patient engagement should therefore account for both interpersonal relationships and team dynamics. Finally, this study illustrates the role of evaluation as an iterative and practice-oriented process. Findings from this evaluation directly informed changes to advisory group engagement, including improvements to onboarding materials, communication strategies, and meeting structures. This reinforces the value of embedding evaluation within research programs as a mechanism for continuous improvement, rather than as a retrospective assessment alone.

Together, these insights contribute to advancing the practice of research evaluation by offering practical considerations for designing, interpreting, and applying evaluations of patient engagement in real-world research settings. This study contributes to research evaluation practice by demonstrating how mixed-methods approaches can be used to generate more nuanced and actionable assessments of patient engagement. By highlighting challenges like interpersonal communication and the importance of team dynamics, these findings support the development of more sensitive and participant-oriented engagement strategies.

### 5.0 Conclusion

This evaluation highlights both the potential and challenges that come with patient engagement, particularly in a large, multicomponent project. POINT study emphasizes the value of flexibility, trust, and opportunities to contribute in ways aligned with patient partners’ capacities. By incorporating member feedback, the project strengthened its engagement process, demonstrating reflective practices and continuous improvement in patient-partnered research. The insights gained through this evaluation contribute to the growing literature on patient-engagement practices and offer guidance for research teams striving to create accessible, equitable, and impactful engagement environments.

## Data Availability

All data produced in the present study are available upon reasonable request to the authors

## Abbreviations

ADHD: Attention-Deficit/Hyperactivity Disorder
POINT: Patient-Oriented Innovative Nutrition Trial
AG: Advisory Group
POR: Patient-Oriented Research
CIHR: Canadian Institutes of Health Research
AbSPORU: Alberta Strategy for Patient-Oriented Research Unit
PORCCH: Patient-Oriented Research Curriculum in Child Health
EDI: Equity, Diversity, and Inclusion
RA: Research Assistant
RC: Research Coordinator
PI: Principal Investigator
PEIRS-22: Patient Engagement in Research Scale (22-item version)
PUFAs: Polyunsaturated fatty acids
MADDY: Micronutrients for Attention-Deficit/Hyperactivity Disorder (ADHD) in Youth

## Ethics approval and consent to participate

The study has been approved by the University of Calgary Conjoint Health Research Ethics Board (REB20-1822). All study participants provided written, electronic, or verbal informed consent to participate.

## Acknowledgements

We gratefully acknowledge the advisory group members of the POINT project for their meaningful contributions. Their engagement in the advisory group, along with their willingness to participate in the survey and interview, contributed substantially to this research. We thank Suzanne Deliscar for reviewing the plain language summary.

## Authors’ contributions

Study concept and design: Kirsten Schmidt, Brenda Leung

Acquisition of data: Sadia Ahmed, Fakhriyya Aghabayli

Data analysis/interpretation: Kirsten Schmidt, Sadia Ahmed, Kaniz Jannat, Brenda Leung

Drafting/preparation of manuscript: Kirsten Schmidt, Sadia Ahmed, Kaniz Jannat

Critical revision: Kirsten Schmidt, Sadia Ahmed, Fakhriyya Aghabayli, Kaniz Jannat, Brenda Leung

## Data availability statement

All summary data are contained within the manuscript and its supplementary files. Data produced in the present study are unavailable for additional analyses. Questions and other requests may be directed to the corresponding authors.

